# FreqFuseNet: Scale-Normalized Dual-Frequency Fusion for Thin-Wall Head-and-Neck OAR Segmentation

**DOI:** 10.64898/2026.07.09.26357642

**Authors:** Wen-Yu Chen, Guan-Yu Lin, Shu-Yen Wan

**Affiliations:** Department of Information Management, Chang Gung University, Taoyuan, Taiwan; Department of Computer Science and Information Engineering, Chang Gung University, Taoyuan, Taiwan

**Keywords:** medical image segmentation, head-and-neck CT, frequency fusion, scale normalization, thin-structure segmentation, false-negative analysis, boundary metrics

## Abstract

Thin-wall organs-at-risk (OARs) in head-and-neck CT are hard to segment because of their small size and boundary sensitivity. FreqFuseNet aligns the activation scales of heterogeneous FFT and FcaNet bottleneck branches before bounded residual fusion; an initialization-time diagnostic indicated a scale mismatch of approximately three orders of magnitude. Under an annotation-defined binary ROI protocol on SegRap2023 (84 training, 18 validation, 18 held-out test patients; ten OARs), the original reporting pipeline gave primary-seed Dice 0.849, HD95 0.824 mm and Surface Dice@1mm 0.959, whereas a native-space standardized re-evaluation gave Dice 0.850 and HD95 1.119 mm for the same seed; these protocols must not be interchanged. Holm-adjusted patient-level tests supported Dice gains over 3D U-Net and MedNeXt-S but no HD95 advantage, the comparison with naive fusion was not significant, and near-zero frequency routing with unchanged masks under frequency-feature zeroing limits frequency-specific attribution. In an exploratory validation cohort, a thickness-conditioned asymmetric objective cut thin-region false negatives by 14.22 percentage points but raised the false-positive rate by 48.01% and worsened surface accuracy, failing its prespecified criteria. No end-to-end localization or clinical generalization is claimed.

## 1 Introduction

Thin-wall head-and-neck organs-at-risk (OARs)—the cochlea, vestibular semicircular canals, internal auditory canal (IAC), tympanic cavity, and middle ear—have wall thicknesses of 0.5–2 mm and volumes of 0.04–1.5 cm^3^, near the resolution limit of clinical CT, so small surface deviations produce large volumetric errors relevant to cochlear- and vestibular-sparing radiotherapy [1].

U-Net-style networks [2, 3] and their successors nnU-Net [4], MedNeXt [5] and SegResNet [6] are strong base-lines for head-and-neck OAR delineation [7, 8], yet small and thin structures remain difficult, and boundary quality is often governed by high-frequency transitions [9]. Frequency-domain features offer a complementary inductive bias [10]: FcaNet [11] extends squeeze-and-excitation attention [12] to DCT bases, GFNet [13] uses global frequency filters, and wavelet-based fusion [14] helps small structures. Our AFS-DSN [15] combined frequency and spatial streams for sinus CT; here we address the less explored incompatibility of activation scales between frequency branches, which within-branch normalization [16, 17] does not resolve. To our knowledge, few studies have characterized this branch-scale incompatibility in FFT–attention dual-frequency fusion under mixed-precision training.

In our implementation, an initialization-time diagnostic showed an activation-scale mismatch of approximately three orders of magnitude between the FFT and FcaNet branches (about 863× in the original diagnostic), so a nominal residual coefficient *β*_*c*_ = 0.05 would correspond to a roughly 43× residual-to-primary magnitude ratio.

FreqFuseNet therefore aligns the FcaNet residual branch to the FFT activation scale before residual injection, with no trainable parameters for the alignment. We evaluated it on SegRap2023 against component variants and external baselines using boundary-sensitive metrics, patient-level statistical testing and two seeds; because the compared architectures differ in more than one design choice, the results characterize complete configurations and do not isolate scale normalization.

## 2 Materials and Methods

### 2.1 Data and ROI Protocol

This is a retrospective secondary analysis of the public SegRap2023 benchmark [7] (head-and-neck CT of nasopharyngeal carcinoma patients, 3.0 mm slices, non-contrast). The 120 official training cases were re-partitioned into one fixed 84/18/18 training/validation/test split used for all models; the official validation and test cases were not used. We study 10 thin-wall OARs (Cochlea, VestibulSemi, IAC, TympanicCavity and MiddleEar, left and right) selected for wall thickness ≤2 mm and radiotherapy relevance [1, 18], giving 18 × 10 = 180 binary per-OAR test samples. Each binary annotation defines a 3D bounding box expanded by 32 voxels per direction; the ROI is resampled to 128^3^, z-score normalized and segmented as background/foreground with softmax and Dice+CE loss. This annotation-defined ROI protocol does not evaluate end-to-end localization.

### 2.2 FreqFuseNet Architecture

Fig. 1 shows the architecture. A U-Net-style encoder [3] (four double-convolution stages with 32–256 channels, max-pooling) maps the ROI to a bottleneck feature **b** ∈ R^512×8×8×8^. The *FFT branch* applies a real 3D FFT [19] to **b**, represents the spectrum by log-compressed magnitude and the cosine and sine of the phase, processes these with independent convolutional blocks, fuses them with a 1 × 1 × 1 convolution, and reconstructs the spectrum with real and imaginary heads before an inverse FFT, giving **F**_FFT_; the heads use small-weight initialization so the output starts near zero. The *FcaNet branch* applies DCT-based multi-spectral channel attention [11] to **b**, giving **F**_FcaNet_.

**Figure 1.**
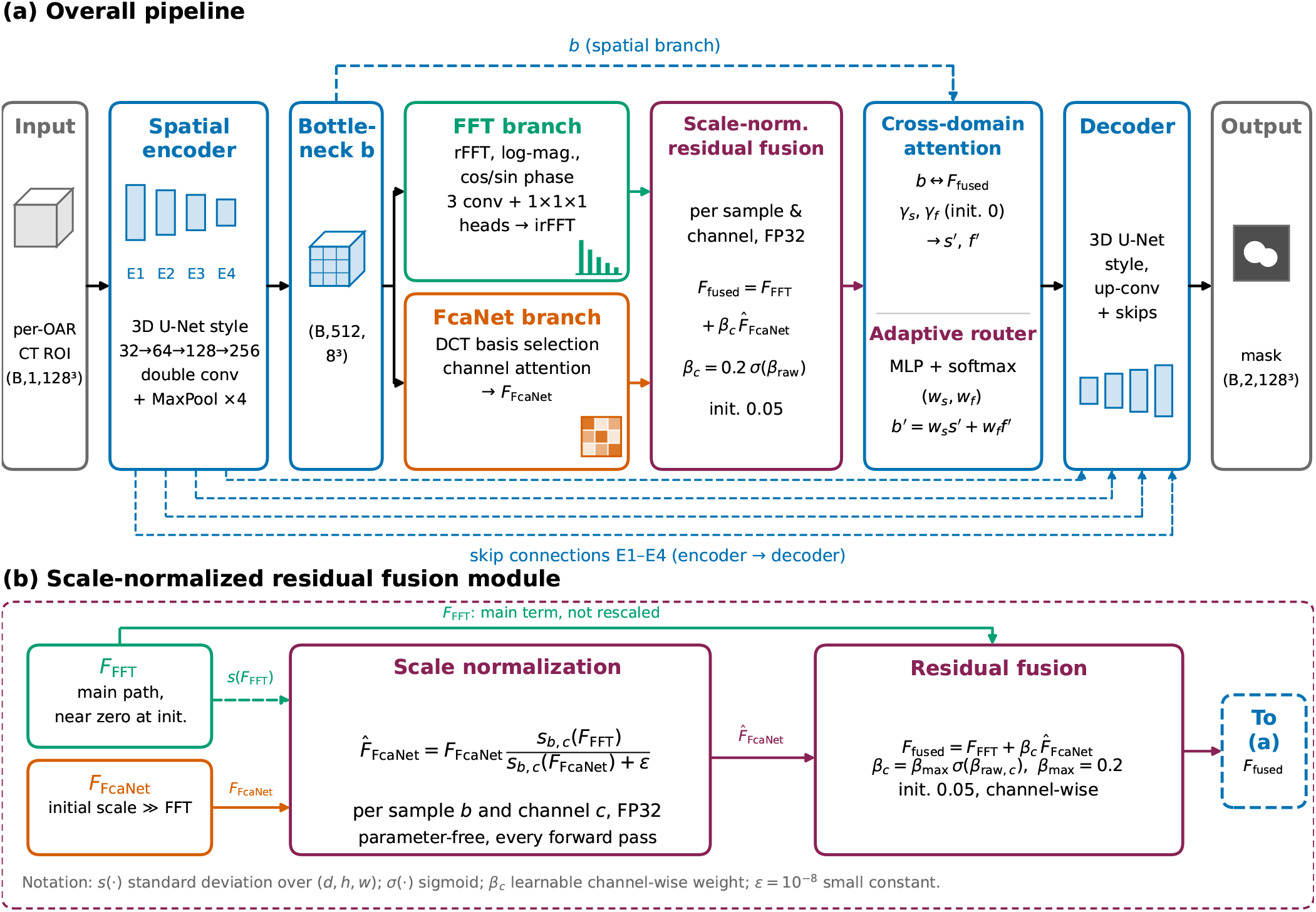
Architecture of FreqFuseNet. (a) A U-Net-style encoder maps the per-OAR CT ROI (*B ×* 1 *×* 128^3^) to the bottleneck **b** (*B ×* 512 *×* 8^3^); an FFT branch and a DCT-based FcaNet branch process **b** in parallel and are combined by scale-normalized residual fusion into **F**_fused_, which enters the cross-domain attention and adaptive router (inherited from AFS-DSN) before decoding into a binary mask (*B ×* 2 *×* 128^3^). Dashed blue lines: direct use of **b** in the cross-domain attention and the encoder–decoder skip connections E1–E4. (b) Fusion module (Eqs. (1)–(2)): the FcaNet output is rescaled to the per-sample, per-channel standard deviation of the FFT output (FP32, parameter-free) and added to it with bounded channel-wise coefficients *β*_*c*_. The diagram shows the designed data flow only; it does not indicate that the frequency pathway contributes to the final predictions (Section 3.1).

#### Scale mismatch

Let *s*_*b,c*_(**F**) = Std_*d,h,w*_(*F*_*b,c,d,h,w*_) for sample *b* and channel *c*, and *r* the ratio of the channel-averaged scales of **F**_FcaNet_ and **F**_FFT_. An initialization-time diagnostic reported *σ*_FFT_ ≈ 0.000679 and *σ*_FcaNet_ ≈ 0.586, i.e. *r* ≈ 863; a separately reported audit with full 128^3^ inputs measured 887–894 at initialization and approximately 865 for the trained seed-2 model, so we describe the mismatch as approximately three orders of magnitude. Naive fusion **F**_FFT_ + *β* **F**_FcaNet_ with *β* = 0.05 would then give a residual-to-primary ratio *βr* ≈ 43, inverting the intended FFT-dominant design. These values characterize the imbalance and are not statistics used in training or inference.

#### Scale-normalized residual fusion

FreqFuseNet rescales the residual branch per sample and channel at every forward pass (FP32, no running statistics, no trainable parameters for the rescaling) and adds it with a bounded coefficient:

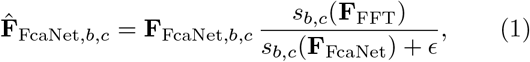

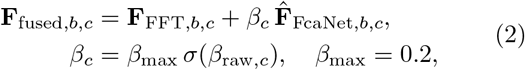

with all *β*_*c*_ initialized at 0.05; after alignment the residual-to-primary scale ratio is approximately *β*_*c*_, i.e. roughly 5% at initialization. Cross-domain attention (learnable scalars *γ*_*s*_, *γ*_*f*_) and the adaptive router (softmax weights (*w*_*s*_, *w*_*f*_) from spatial statistics and frequency-band en-ergies, **b**′ = *w*_*s*_**s**′ + *w*_*f*_ **f**′) are inherited unchanged from AFS-DSN [15]; the contribution of this work is the scale-normalized FFT-main residual branch.

### 2.3 Training, Ablation Arms and Baselines

Models were trained with Dice+cross-entropy loss and AdamW [20] (peak learning rate 1 × 10^−4^, weight decay 1 × 10^−5^), five warm-up epochs and cosine decay [21] over 100 epochs, mixed precision [22] with the FFT and scale-normalization operations in FP32, and the checkpoint with the highest validation Dice, on one NVIDIA A40 GPU. Internal models used batch size 2 and external baselines batch size 1. FreqFuseNet was trained with seeds 2 and 3; seed 2 is the primary seed for patient-level comparisons. It has 29.7 M parameters, 92.8% fewer than DWT-Full (414.6 M).

Ablation arms: Stage 1, AFS-DSN baselines with a full DWT encoder (DWT-Full), a lightweight one (DWT-Lite, 27.4 M) or focal frequency loss (*λ* ∈ {0.05, 0.10, 0.20}); Stage 2, FcaNet (24.0 M); Stage 3, FFT (29.4 M); Stage 5B, FixedFusion, a learned channel-wise convex mixture of both branches (29.7 M, FFT weight initialized at 0.5); Stage 5C, FreqFuseNet. Stage 5C differs from 5B in fusion formula, coefficient range, initialization and scale alignment. External baselines were 3D U-Net (19.2 M), MedNeXt-S (5.6 M) and SegResNet (18.8 M), trained from scratch under the same split, ROIs, loss, schedule, checkpoint criterion and evaluation. A standard nnU-Net is excluded because its pipeline targets whole-volume multi-class segmentation.

### 2.4 Evaluation Protocols and Statistics

Three protocols must be distinguished: (i) the *historical ROI-resampled reporting pipeline* (Table 1); (ii) a *standardized surface-distance recomputation on the* 128^3^ *grids* (Fig. 2); and (iii) a *native-space standardized re-evaluation* (Section 3.2). They can give materially different HD95 values and rankings and are not interchangeable. We report Dice, HD95 and Surface Dice at 1 and 2 mm [18], with paired inference over the *18 patients*, not 180 independent observations. Historical Wilcoxon analyses [23] were uncorrected and are not confirmatory. The native-space audit uses a family of 12 patient-paired comparisons (exact Wilcoxon signed-rank, FreqFuseNet seed 2 versus three external baselines on four metrics) with Holm correction [24]; its primary analysis capped the single non-finite MedNeXt-S HD95 value at the ROI diagonal. These audit statistics have not been independently reproduced within this submission package.

**Table 1.** Ablation and baseline results on 10 thin-wall OARs (SegRap2023, 180 binary per-OAR test samples from 18 test cases). Historical ROI-resampled metric pipeline; not comparable with the native-space values in the text or with Fig. 2. Bold marks historical best values, not corrected rankings.

| Method | Params (M) | Dice | HD95 (mm) | SDice@1mm | SDice@2mm |
|---|---|---|---|---|---|
| *External baselines* |  |  |  |  |  |
| 3D U-Net | 19.2 | 0.830 | 1.002 | 0.945 | 0.976 |
| MedNeXt-S | 5.6 | 0.825 | 0.948 | 0.945 | 0.978 |
| SegResNet | 18.8 | 0.846 | 0.848 | 0.956 | 0.983 |
| *Ablation stages* |  |  |  |  |  |
| DWT-Full (S1) | 414.6 | 0.840 | 0.884 | 0.952 | 0.984 |
| DWT-Lite (S1) | 27.4 | 0.838 | 1.030 | 0.949 | 0.977 |
| FFL *λ* = 0.05 (S1) | 414.6 | 0.841 | 0.904 | 0.955 | 0.983 |
| FFL *λ* = 0.10 (S1) | 414.6 | 0.839 | 0.879 | 0.952 | 0.984 |
| FFL *λ* = 0.20 (S1) | 414.6 | 0.838 | 0.867 | 0.951 | 0.983 |
| FcaNet (S2) | 24.0 | 0.842 | 0.837 | 0.956 | 0.983 |
| FFT (S3) | 29.4 | 0.840 | 0.841 | 0.957 | 0.986 |
| FixedFusion (S5B) | 29.7 | 0.843 | 0.871 | 0.955 | 0.984 |
| FreqFuseNet (seed 2) | 29.7 | **0.849** | 0.824 | **0.959** | **0.987** |
| FreqFuseNet (seed 3) | 29.7 | 0.843 | **0.822** | 0.957 | 0.986 |

**Figure 2.**
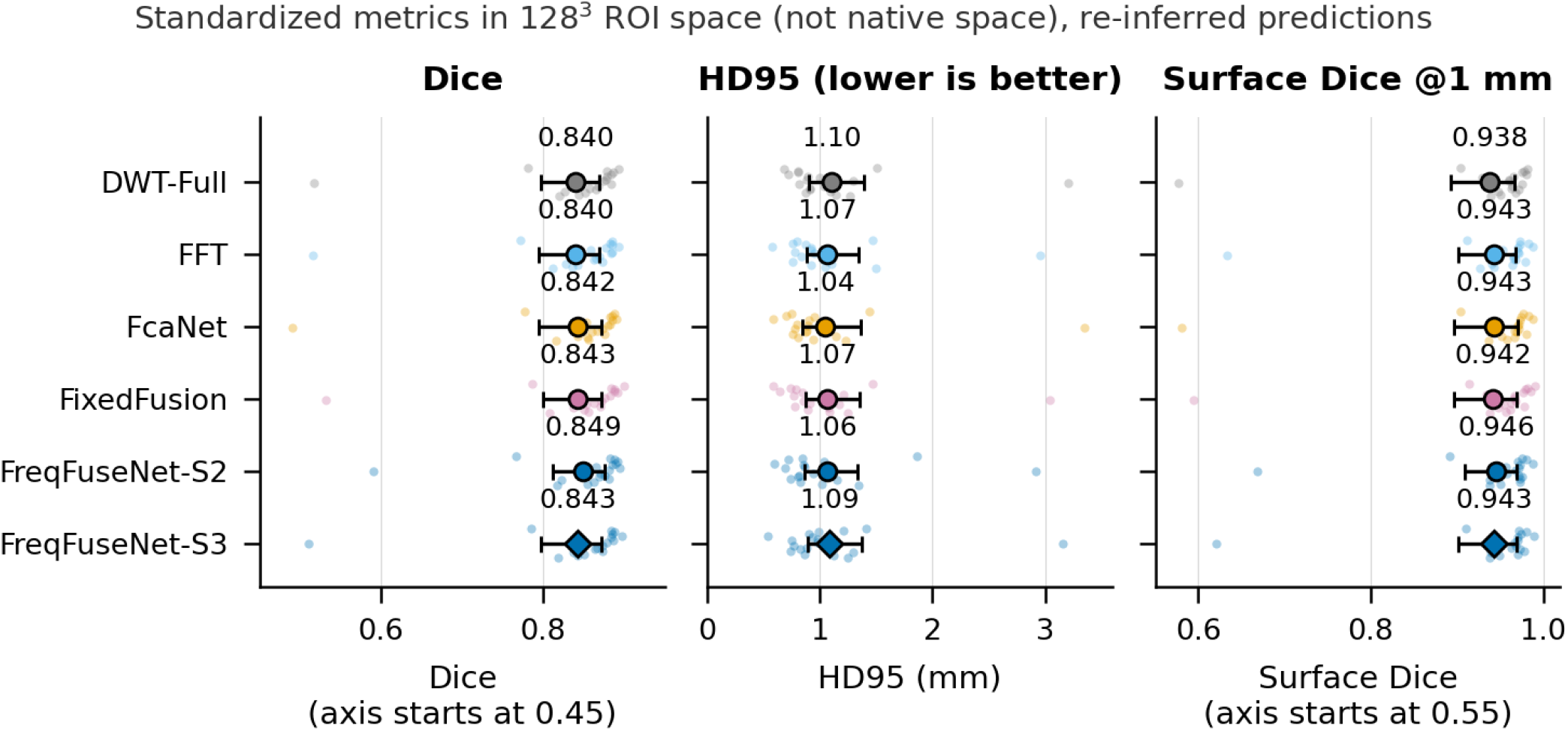
Ablation under standardized metrics in 128^3^ ROI space (not native space): Dice, standardized HD95 and Surface Dice at 1 mm, with per-ROI voxel spacing and predictions re-inferred from the same checkpoints. Test cohort (18 cases, 180 case–OAR samples); all methods share cases, spacing and metric code. Markers: mean with 95% case-bootstrap intervals; dots: per-case values; Dice and Surface Dice axes do not start at zero. HD95 here is not comparable with Table 1 or the native-space values. After Holm correction (36 exploratory tests; smallest adjusted *p* = 0.136) no paired difference between FreqFuseNet (seed 2) and the other arms is significant.

## 3 Results

### 3.1 Scale Mismatch, Frequency Routing and Co-efficients

After normalization the designed residual-to-primary ratio is 0.05, a property of the fusion block before adaptive routing and not a measurement of the trained models’ effective frequency contribution. In Stage 5B the learned FFT fusion weight stayed near its 0.5 initialization (0.5009 ± 0.0016). An audit of the two frozen Stage 5C checkpoints found median router frequency weights of approximately 2.53 × 10^−6^ (seed 2) and 3.01 × 10^−4^ (seed 3); setting the routed frequency-fusion feature to zero at inference for 180 per-OAR observations per seed changed the final masks in *zero* voxels. This tests the frequency feature at the routing point, not every route by which frequency-derived statistics could act through cross-attention, routing or training; it shows insensitivity of the final masks, not that the frequency module was unnecessary during training, so the initialization-scale diagnostic is not proof of frequency-specific gains. The residual coefficients also stayed near 0.05 (all 512 seed-2 raw coefficients identical to their float32 initialization; seed-3 mean raw shift approximately 8.54 × 10^−6^); short gradient probes on real training ROIs gave raw-coefficient gradients of 10^−12^–10^−13^ with scale normalization versus 10^−9^–10^−10^ without, consistent with update attenuation under the AdamW *ϵ* = 10^−8^, and characterize optimization behavior, not segmentation efficacy.

### 3.2 Segmentation Performance

Under the historical ROI-resampled protocol (Table 1), FreqFuseNet achieved Dice 0.849, HD95 0.824 mm, SDice@1mm 0.959 and SDice@2mm 0.987 in the primary seed, and 0.843, 0.822 mm, 0.957 and 0.986 in the second seed. Compared with FixedFusion, mean HD95 improved from 0.871 to 0.824 mm, but the paired difference was not significant (Wilcoxon *p* = 0.054; Dice *p* = 0.181, SDice@1mm *p* = 0.580, SDice@2mm *p* = 0.212), and the comparison does not isolate scale normalization. Seed-2 Dice was 0.849 versus 0.830 (3D U-Net), 0.825 (MedNeXt-S) and 0.846 (SegResNet); the unadjusted historical tests (Dice *p* = 0.000191 and 0.000107 versus 3D U-Net and MedNeXt-S) are not multiplicity-adjusted evidence. In the native-space audit, Holm-adjusted Dice *p* = 0.00092 versus 3D U-Net and *p* = 0.00210 versus MedNeXt-S, with a marginal SDice@1mm difference versus MedNeXt-S (adjusted *p* = 0.0475); no significant Dice difference from SegResNet and no Holm-significant HD95 advantage were established, so uniform boundary superiority is not claimed.

The native-space summaries (transcribed from the research handoff; not comparable with Table 1) give Dice/HD95 of 0.823/1.338 mm (3D U-Net), 0.848/1.124 mm (SegResNet), 0.846/1.131 mm (Fixed-Fusion), 0.850/1.119 mm (FreqFuseNet seed 2) and 0.843/1.103 mm (seed 3), and an HD95 of 1.0846 mm for FcaNet, lower than seed 2. MedNeXt-S has no finite, comparably aggregated native-space HD95 because one case was non-finite, and no case was excluded for convenience; unreported values were not inferred. A third protocol, standardized HD95 on the resampled 128^3^ grid, gave approximately 1.063 mm (seed 2), 1.088 mm (seed 3), 1.042 mm (FcaNet) and 1.071 mm (SegResNet) and must not be substituted for native-space numbers. Source arrays and metric-pipeline versions still require reconciliation before these audit statistics can be regarded as independently reproduced.

### 3.3 Component Ablation and Per-OAR Results

Under the standardized evaluation in 128^3^ ROI space (not native space; predictions re-inferred from the same check-points, same 18 test cases and per-ROI spacing; Fig. 2), mean Dice was 0.849 for FreqFuseNet seed 2 versus 0.840– 0.843 for the other arms, mean HD95 1.063 mm versus 1.042–1.101 mm (FcaNet lowest, DWT-Full highest), and mean Surface Dice@1mm 0.946 versus 0.938–0.943. The bootstrap intervals overlap substantially, and after Holm correction across 36 exploratory comparisons no paired difference remained significant (smallest adjusted *p* = 0.136, Dice versus FFT; unadjusted *p* = 0.004), so FreqFuseNet has the highest mean Dice but no distinguishable HD95 advantage under this protocol. In the historical comparison (Table 1), FcaNet (24 M) improved HD95 by 0.047 mm and SDice@1mm by 0.004 over DWT-Full (414.6 M), and DWT-Lite degraded HD95 by 0.146 mm, so differences are not explained by parameter count alone. Relative to DWT-Full under the historical ROI definition, Dice improved on all 10 OARs (largest: TympanicCavity L, +0.015; IAC L, +0.014) and HD95 on 8 of 10 (TympanicCavity L/R, −0.202/ −0.207 mm); the surface-distance ranking needs reassessment under the native-space protocol, and per-OAR tests are exploratory. Fig. 3 shows one deliberately chosen test slice.

**Figure 3.**
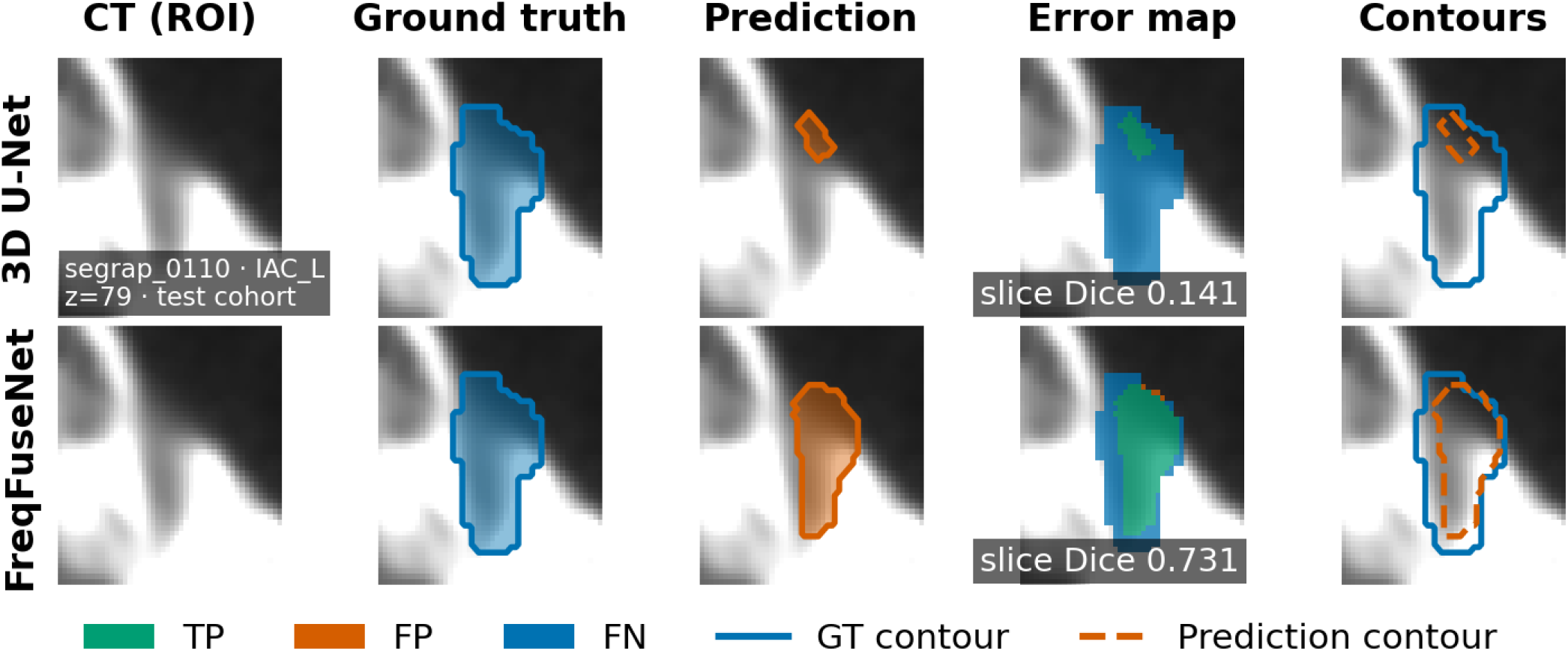
One deliberately chosen axial slice (index 79 of the 128^3^ ROI) of IAC_L in segrap_0110 (test cohort) for 3D U-Net (top) and FreqFuseNet (bottom): CT, ground truth, prediction, error map (TP, FP, FN) and contours. Slice Dice from the displayed arrays is 0.141 (3D U-Net) and 0.731 (FreqFuseNet); the 3-D ROI Dice is 0.289 and 0.661, and on the adjacent slice with identical ground-truth area the slice Dice is 0.814 and 0.847. It is not evidence of a frequency-specific mechanism or of a cohort-wide advantage.

### 3.4 Exploratory Validation-Cohort Analyses

These analyses used the 18-patient *validation* cohort, which was inspected repeatedly during development; they are not independent confirmation (Fig. 3 above is a single illustrative test-cohort slice and is likewise not evidence of an average effect). Foreground false negatives in regions thinner than 2 mm (a distance-transform proxy, not an anatomical measurement) were 24.5%, versus 4.2% for 2–4 mm and 2.5% for ≥ 4 mm, an observational pattern confounded by organ size and spacing. Forward label re-sampling with legacy nearest interpolation gave a −22.3% mean physical-volume difference across 180 observations, a property of that discretization and not proof of information loss (an earlier round-trip Dice of 0.401 was invalid; center-aligned reconstruction gave 0.969). A boundary-conditioned frequency residual (BCFRF) changed Dice by +0.00057 and HD95 by −0.0275 mm relative to a spatial-residual control (4,291 boundary corrections versus 5,177 new errors) and was discontinued.

Thickness-conditioned asymmetric learning (TCAL) was compared in three matched 500-step fine-tuning arms from the seed-2 checkpoint: A, cross-entropy plus Dice; B, local-thickness-weighted; C, B plus penalties on thin-region false negatives and background false positives. The primary endpoint was the thin-region false-negative rate of the four target OARs (bilateral TympanicCavity and MiddleEar), C versus B, with 10,000 patient-level boot-strap resamples (seed 20261031); prespecified criteria were ≥ 5 points false-negative reduction, ≤ 2 points Dice regression, ≤ 5% relative false-positive increase and no material HD95 or Surface Dice deterioration. Arm C lowered thin-region false negatives from 30.08% to 15.86% (−14.22 points, 95% CI [−15.90, −12.59]; 18/18 patients) but raised the background false-positive rate by 48.01%, lowered Dice from 0.7646 to 0.7531, raised HD95 by 0.180 mm and lowered Surface Dice@1mm by 0.0140, so it *failed* the false-positive and boundary safe-guards. These metrics come from a frozen evaluator whose HD95 implementation differs from the standardized code in an unresolved way, so TCAL HD95 is protocol-specific and not comparable with other HD95 values here; three patients with patient-mean HD95 of 8–10 mm were valid observations and retained. The result is a recall– oversegmentation trade-off, not an improvement.

## 4 Discussion

FreqFuseNet targets a specific problem: a large activation-scale mismatch can make the effective contribution of a residual branch differ sharply from its nominal coefficient, and aligning the FcaNet feature to the FFT branch at every forward pass fixes this relationship without extra parameters. Under the annotation-defined ROI protocol the complete architecture achieved competitive overlap and boundary metrics, with Holm-adjusted significant Dice gains over 3D U-Net and MedNeXt-S but no Holm-significant HD95 advantage, only a marginal SDice@1mm advantage over MedNeXt-S (adjusted *p* = 0.0475), and no distinguishable HD95 under the standardized 128^3^ comparison. Because FixedFusion differs in more than normalization, the results support the full architecture without assigning the improvement to scale alignment, and the near-zero frequency routing, the unchanged masks under zeroing and the negative BCFRF result weaken any claim of frequency-specific boundary correction; causal attribution would need matched normalization and branch-removal interventions. TCAL lowers thin-region omission at the cost of false positives and surface accuracy; since arm C failed its safeguards, overlaps with established loss families and was assessed on a reused validation cohort, it is not presented as an improvement or a novel loss.

### Limitations

Evaluation used 18 test cases and two seeds; Stages 5B and 5C are confounded beyond normalization, and the coefficient probes were not matched across settings. ROIs are annotation-derived (no whole-volume localization), no standard nnU-Net was bench-marked, and external baselines used batch size 1 versus 2 with one seed. The audit statistics need reconciliation of source arrays and pipeline versions, and the three metric protocols must not be conflated. Sex, gender and age were not analyzed. The exploratory experiments used one seed and a reused validation cohort, and Fig. 3 is a single slice. Distances must be read against the 3.0-mm slice thickness; external validation, dosimetry and prospective testing remain future work.

## 5 Conclusions

FreqFuseNet aligns the FcaNet residual to the FFT primary branch before fusion, after an activation-scale mismatch of about three orders of magnitude was identified at initialization. Under the historical protocol seed 2 gave Dice 0.849, HD95 0.824 mm and SDice@1mm 0.959 (native-space audit: Dice 0.850, HD95 1.119 mm). The adjusted audit supports Dice gains over 3D U-Net and MedNeXt-S but not uniform surface-distance superiority, and the routing and exploratory results do not support frequency-specific attribution or an overall improvement. Independent evaluation and targeted causal ablations remain necessary.

## Data Availability

All data analyzed in this study are derived from the publicly available SegRap2023 head-and-neck CT dataset (https://segrap2023.grand-challenge.org). Processed data and trained models used in this work are available from the corresponding author upon reasonable request.

## Ethics approval and consent to participate

This secondary analysis used only the publicly available, de-identified SegRap2023 data; no participants were recruited, no additional ethics approval was obtained, and consent is not applicable.

## Data availability

The SegRap2023 dataset is publicly available through the official Grand Challenge platform at https://segrap2023.grand-challenge.org/dataset/; access may require registration and acceptance of the platform’s terms.

## Code availability

The code supporting the findings of this study is available from the corresponding author upon reasonable request.

## CRediT authorship contribution statement

**Wen-Yu Chen:** Conceptualization, Methodology, Software, Validation, Formal analysis, Investigation, Data curation, Writing – original draft, Writing – review and editing, Visualization. **Guan-Yu Lin:** Methodology, Software, Validation, Formal analysis, Investigation, Data curation, Writing – original draft, Writing – review and editing, Visualization. **Shu-Yen Wan:** Supervision, Project administration, Funding acquisition.

## Acknowledgements

The authors thank the Medical Imaging Laboratory at Chang Gung University, RunPod for computational resources, and the creators of SegRap2023 for open-access data.

